# Prolonged low-dose methylprednisolone in patients with severe COVID-19 pneumonia

**DOI:** 10.1101/2020.06.17.20134031

**Authors:** Francesco Salton, Paola Confalonieri, Pierachille Santus, Sergio Harari, Raffaele Scala, Simone Lanini, Valentina Vertui, Tiberio Oggionni, Antonella Caminati, Vincenzo Patruno, Mario Tamburrini, Alessandro Scartabellati, Mara Parati, Massimiliano Villani, Dejan Radovanovic, Sara Tomassetti, Claudia Ravaglia, Venerino Poletti, Andrea Vianello, Anna Talia Gaccione, Luca Guidelli, Rita Raccanelli, Paolo Lucernoni, Donato Lacedonia, Maria Pia Foschino Barbaro, Stefano Centanni, Michele Mondoni, Matteo Davì, Alberto Fantin, Xueyuan Cao, Lucio Torelli, Antonella Zucchetto, Marcella Montico, Annalisa Casarin, Micaela Romagnoli, Stefano Gasparini, Martina Bonifazi, Pierlanfranco D’Agaro, Alessandro Marcello, Danilo Licastro, Barbara Ruaro, Maria Concetta Volpe, Reba Umberger, Umberto Meduri, Marco Confalonieri

## Abstract

**Background:** In hospitalized patients with COVID-19 pneumonia, progression to acute respiratory failure requiring invasive mechanical ventilation (MV) is associated with significant morbidity and mortality. Severe dysregulated systemic inflammation is the putative mechanism. We hypothesize that early prolonged methylprednisolone (MP) treatment could accelerate disease resolution, decreasing the need for ICU and mortality.

**Methods:** We conducted a multicenter, observational study to explore the association between exposure to prolonged, low-dose, MP treatment and need for ICU referral, intubation or death within 28 days (composite primary endpoint) in patients with severe COVID-19 pneumonia admitted to Italian respiratory high-dependency units. Secondary outcomes were invasive MV-free days and changes in C-reactive protein (CRP) levels.

**Results:** Findings are reported as MP (n=83) vs. control (n=90). The composite primary endpoint was met by 19 vs. 40 [adjusted hazard ratio (HR) 0.41; 95% confidence interval (CI): 0.24-0.72]. Transfer to ICU and need for invasive MV was necessary in 15 vs. 27 (p=0.07) and 14 vs. 26 (p=0.10), respectively. By day 28, the MP group had fewer deaths (6 vs. 21, adjusted HR=0.29; 95% CI: 0.12-0.73) and more days off invasive MV (24.0 ± 9.0 vs. 17.5 ± 12.8; p=0.001). Study treatment was associated with rapid improvement in PaO_2_:FiO_2_ and CRP levels. The complication rate was similar for the two groups (p=0.84).

**Conclusion:** In patients with severe COVID-19 pneumonia, early administration of prolonged MP treatment was associated with a significantly lower hazard of death (71%) and decreased ventilator dependence. Randomized controlled studies are needed to confirm these findings.

**Registration:** ClinicalTrials.gov. Identifier: NCT04323592

## Introduction

Italy was the first European Country overwhelmed by the SARS-CoV-2 pandemic, experiencing an unsustainable burden on the healthcare system. The greatest impact was on intensive care units (ICUs) because 16% of hospitalized cases developed acute respiratory failure (ARF) requiring ICU admission.[1] COVID-19 patients with ARF necessitate weeks of mechanical ventilation (MV) and have an unacceptably high mortality rate.[2] This is an unprecedented global emergency where even countries with advanced health care systems rapidly reach ICU saturation, and intensivists are forced to make difficult ethical decisions that are uncommon outside war zones. Any intervention directed at decreasing dependence on ventilators and mortality in COVID-19 patients is an ethical imperative and would have a significant global impact on public health.

Over the last few decades, Italy has built-up a diffuse network of respiratory high dependency units (RHDUs) which also treat patients with severe pneumonia-related ARF requiring continuous monitoring and noninvasive positive pressure ventilation (NPPV).[3] Patients with disease progression who require endotracheal intubation are transferred to the ICU. During the pandemic, RHDUs were pivotal in reducing ICU referral.[4]

Indeed, patients with severe COVID-19 have exhausted antiviral defenses and massive tissue and systemic inflammatory response. Corticosteroids are powerful anti-inflammatory drugs that could have a role in promoting the resolution of ARF in patients with severe COVID-19 infection.[5] The rationale for prolonged, low-dose, corticosteroid treatment in severe COVID-19 was recently reviewed.[6]

We hypothesized that early MP treatment in patients with severe SARS-CoV-2 pneumonia at higher risk for ARF progression requiring invasive MV, may quicken disease resolution, reducing the need for ICU support and mortality. We investigated the association between early intervention with prolonged MP treatment in this high-risk group and the risk for ICU admission, the need for invasive MV or all-cause death by day 28.

## Methods

### Study design, setting and participants

We conducted a multicenter, observational, longitudinal study to evaluate the association between MP treatment and outcome in consecutive patients with severe COVID-19 pneumonia admitted to fourteen Italian RHDUs between February 27^th^ and April 24^th^, 2020. Follow-up continued through May 21^st^, 2020. The composite primary endpoint included admission to ICU, need for invasive MV, or all-cause death by day 28, while secondary endpoints were MV (invasive or noninvasive)-free days by day 28[7] and changes in C-reactive protein (CRP) levels. The study was carried out in accordance with the Declaration of Helsinki. It was registered on Clinicaltrials.gov (NCT043235929) after approval by the referral Ethics Committee for the Coordinating Centre (University Hospital of Trieste, #CEUR-2020-Os-052).

Study baseline was defined as the time of inclusion criteria fulfillment after admission to RHDU. Inclusion criteria were the followings: 1) SARS-CoV-2 positive (on swab or bronchial wash); 2) age >18 years and <80 years; 3) PaO_2_:FiO_2_ <250 mmHg; 4) bilateral infiltrates; 5) CRP >100 mg/L; and/or 6) diagnosis of acute respiratory distress syndrome (ARDS) according to the Berlin definition[8] as an alternative to criteria 4) and 5). Exclusion criteria were: heart failure as main cause of ARF, decompensated liver cirrhosis, immunosuppression (i.e. cancer on treatment, post-organ transplantation, HIV-positive, on immunosuppressant therapy), dialysis-dependence, on long-term oxygen or home mechanical ventilation, idiopathic pulmonary fibrosis, neuromuscular disorders, dementia or a decompensated psychiatric disorder, severe neurodegenerative conditions, on chronic steroid therapy, pregnancy, a do-not-resuscitate order, and use of Tocilizumab or other experimental treatment. Patients in both study groups received standard of care, comprising noninvasive respiratory support, antibiotics, antivirals, vasopressors, and renal replacement therapy as deemed suitable by the healthcare team.

Exposure to methylprednisolone (non-patented drug, ATC code H02AB04) complied with the following protocol: a loading dose of 80 mg iv at study entry (baseline), followed by an infusion of 80 mg/day in 240 mL normal saline at 10 mL/h for at least eight days, until achieving either a PaO_2_:FiO_2_ > 350 mmHg or a CRP < 20 mg/L. After which, oral administration at 16 mg or 20 mg iv twice daily until CRP reached < 20% of normal range or a PaO_2_:FiO_2_ > 400 (alternative SatHbO_2_ ≥ 95% on room air). The MP protocol was developed by the coordinating Center in accordance with the “recommendation for COVID-19 clinical management” by the National Institute for the Infectious Diseases “L. Spallanzani”, Rome.[9] The decision to apply the protocol to COVID-19 was left to the discretion of the treating team for each individual patient. Unexposed patients (controls) were selected from concurrent consecutive COVID-19 patients with the same inclusion and exclusion criteria.

### Data sources and variables

Demographic details, laboratory, clinical and outcome variables were manually extracted from electronic medical records or charts and anonymously coded onto in a standardized data collection form. Three independent physicians checked the data and two researchers adjudicated any difference in interpretation between the primary reviewers.

Serial measurements included: arterial blood gas, CRP, D-dimer, white cell count with differential, hemoglobin, variables for the calculation of the SOFA score,[10] days free from invasive or non-invasive MV until study day 28. Laboratory methodologies, including SARS-CoV-2 detection by reverse-transcriptase polymerase chain reaction (RT-PCR) and reference values were comparable between centers. Other collected data included: date of death, admission to ICU, dates of discharge from hospital and ICU, intra-hospital medications, in-hospital adverse events and comorbidities. Samples from seriated nasopharyngeal swabs were collected in each group to evaluate viral shedding.

### Statistical methods

Considering a study power (1-beta) of 80% and a probability of type 1 error (alpha) of 0.05, assuming that the proportion of treated patients having the primary endpoint was 0.7 under the null hypothesis (according to available information from Fang et al. 2020[11]) and 0.42 under the alternative hypothesis, and considering a 5% dropout rate, a minimum study sample of 104 patients was established. Data were described using absolute and relative frequencies (percentage) or position indices (mean or median) and relative dispersion indices (standard deviation or interquartile range), as appropriate according to the type and distribution of the variable analyzed. The differences between study groups (MP-treated and control) in the proportion of patients reaching the primary endpoint was evaluated by a two-sided Chi-square test. The difference in numerical variables between groups was calculated using Student’s T-test or Wilcoxon rank-sum test, depending on the distribution of the variables.

Differences between study groups concerning categorical or dichotomous variables were evaluated by means of the Chi-square test or Fisher’s exact test, as appropriate. Time-to-event analyses were performed for both the composite primary endpoint and death alone. Time at risk for all-cause death was computed from the date of study enrollment up to the date of death, hospital discharge, or 28 days, whichever came first. Event-free probabilities were estimated by the Kaplan-Meier method and differences between groups were assessed by the log-rank test. Multivariable Cox proportional-hazard models estimated the hazard ratio (HR) of both the primary composite endpoint and all-cause death, with the corresponding 95% confidence intervals (95% CI), taking into account the confounding factors (i.e., sex, age, and baseline values of SOFA score, PaO_2_:FiO_2_, CRP levels) potentially associated with the outcome. These variables and others with baseline differences (e.g. smoke) were tested in univariate survival models and variables significant at p=0.1 were tested in the multivariable model using stepwise selection. Proportional hazards assumption was assessed by visual inspection of the log(-log(survival)) plot. There were no missing data with regard neither to the composite primary endpoint and the adjustment factors included in the final Cox models, nor to MV-free days. Available case analysis was performed for time variation of C-reactive protein (CRP) and PaO_2_:FiO_2_ levels. All tests were two-sided and a p-value of <0.05 was considered as statistically significant.

Sensitivity analyses were completed as recommended by STROBE guidelines for reporting observational studies.[12] Although a protocol was used to standardize study measures, we conducted a sensitivity analysis to account for potential variance in medical decision making that could potentially impact the primary composite outcome. We examined hypothetical scenarios against the hypothesis by varying the number of subjects meeting the primary composite outcome by 3 and 5 subjects to account for potential bias in both groups.

## Results

Between February 27^th^ and April 24^th^, 2020, 322 consecutive SARS-CoV-2-positive patients who were admitted to one of 14 RHDUs with severe pneumonia, were assessed for study eligibility. A total of 173 patients (83 MP-treated exposed and 90 untreated controls) were enrolled, while 149 were excluded as detailed in **Figure 1**.

**Figure 1.**
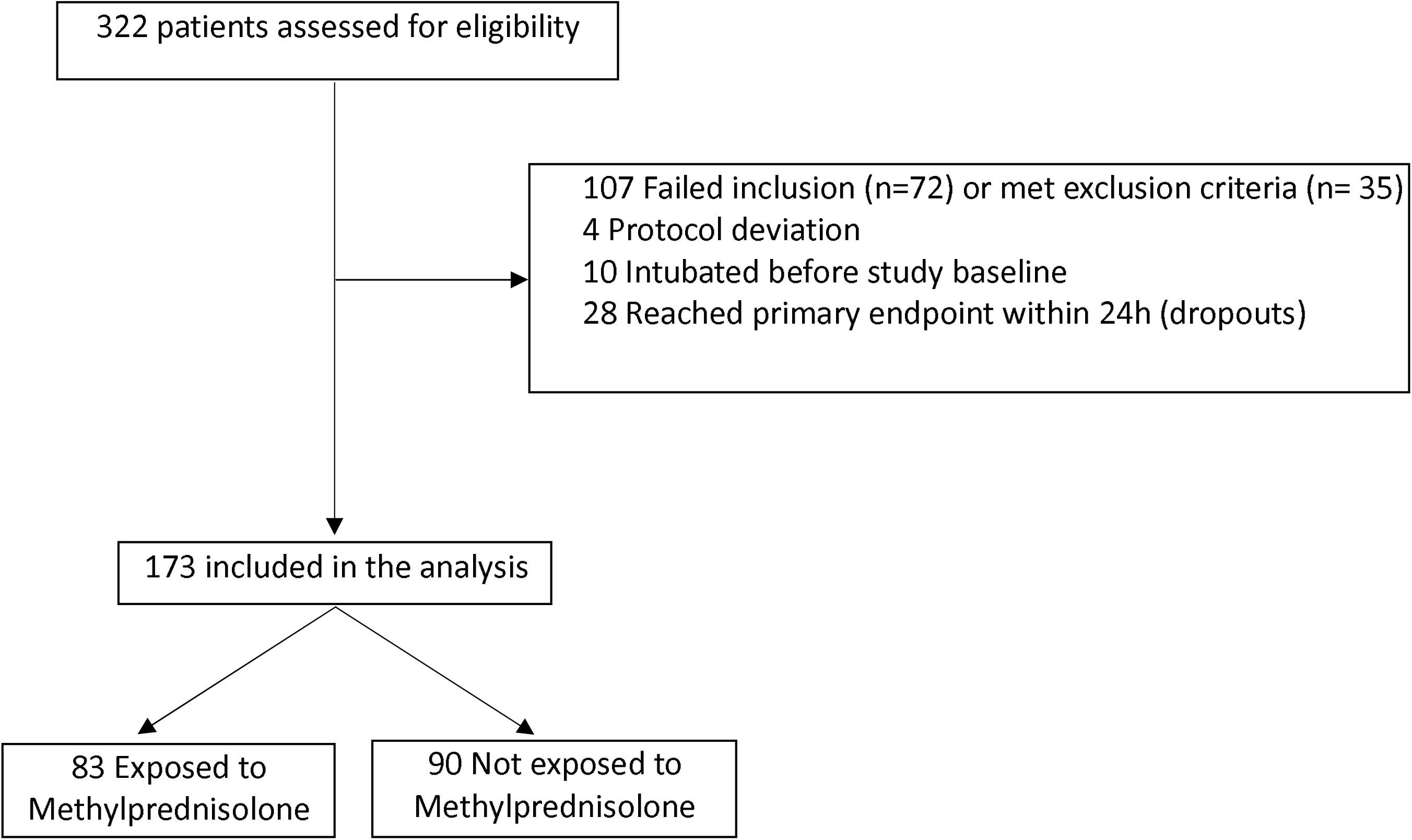
Flow-chart of the study population. Failed to meet inclusion criteria (n=72): age > 80 years old (n=9), criteria for PaO_2_:FiO_2_, C-reactive protein level, or ARDS (n=63). Met exclusion criteria (n=35): heart failure as main cause of ARF (n=2), decompensated liver cirrhosis (n=3), on long-term oxygen therapy and/or home ventilation (n=2), dementia or severe neurodegenerative condition (n=14), active cancer (n=3), on chronic steroid therapy (n=4), use of Tocilizumab or other experimental treatment (n=7). 28 patients who reached the primary endpoint before admission to RHDU or within 24 hours from admission to RHDU were excluded from the analysis; 20 out of these 28 patients did not start MP treatment.

Findings are reported as MP group vs. control group. RHDU admission days to study enrollment were comparable (0.83 ± 2.02 vs. 0.56 ± 1.50, p=0.32). **Table 1** shows how the patients’ baseline characteristics did not differ between groups. The mean duration of MP treatment was 9.11 ± 2.4 days. **Table 2** reports the main study outcomes. The composite primary endpoint was reached by 19 vs. 40 (22.9% vs 44.4%, p=0.003) [adjusted hazard ratio (HR) 0.41; 95% confidence interval (CI): 0.24-0.72] indicating a reduction of 59% in the risk of ICU referral, invasive MV, or death within 28 days. In particular, ICU transfer was necessary in 15 vs. 27 (18.1% vs 30.0%, p=0.07) and for invasive MV in 14 vs. 26 (16.9% vs. 28.9%, p = 0.10).

**Table 1:** Distribution of 173 study patients according to study group and baseline characteristics.

|  | **Methylprednisolone**<br>**(N=83)** | **Control**<br>**(N=90)** | **p-value<sup>o</sup>** |
| --- | --- | --- | --- |
| Age, mean (SD) | 64.4 (10.7) | 67.1 (8.2) | 0.07 |
| Male sex, no. (%) | 54 (65.1) | 66 (73.3) | 0.25 |
| BMI $\geq 30$ kg/m<sup>2</sup>, no. (%)<sup>†</sup> | 19 (33.3) | 18 (32.7) | 1.00 |
| Ever smoker, no. (%)<sup>□</sup> | 22 (29.7) | 29 (45.3) | 0.05 |
| Presence of major co-morbidities, no. (%) | 63 (75.9) | 74 (82.2) | 0.35 |
| Hypertension, no. (%) | 36 (43.4) | 51 (56.7) | 0.09 |
| Diabetes, no. (%) | 19 (22.9) | 25 (27.8) | 0.49 |
| Asthma/COPD, no. (%) | 7 (8.4) | 9 (10.0) | 0.80 |
| OSAS/OHS, no. (%) | 5 (6.0) | 7 (7.8) | 0.77 |
| Congestive heart failure, no. (%) | 4 (4.8) | 2 (2.2) | 0.43 |
| Ischemic cardiovascular disease, no. (%) | 2 (2.4) | 9 (10.0) | 0.06 |
| Chronic kidney disease, no. (%) | 5 (6.0) | 4 (4.4) | 0.74 |
| History of malignancy, no. (%) | 7 (8.4) | 4 (4.4) | 0.36 |
| PaO<sub>2</sub>:FiO<sub>2</sub>, mmHg, mean (SD) | 152.0 (49.8) | 151.0 (60.3) | 0.90 |
| Respiratory rate, breaths/minute, mean (SD)<sup>§</sup> | 23.7 (5.9) | 25.3 (6.8) | 0.16 |
| CRP, mg/L, mean (SD) | 136.9 (72.6) | 148.6 (75.6) | 0.30 |
| D-dimer, ug/FEU/L, median (IQR) | 780 (540-1214) | 871 (472-1517) | 0.82 |
| LDH, U/L, mean (SD) | 370.5 (130.9) | 395.3 (169.3) | 0.34 |
| Lymphocyte count, mean (SD) | 916.2 (657.0) | 954.5 (914.7) | 0.76 |
| SOFA score, median (IQR) | 3 (2-4) | 3 (2-4) | 0.96 |

Legend: SD, standard deviation; IQR inter-quartile range; CRP, C-reactive protein; SOFA, Sequential Organ Failure Assessment; PaO<sub>2</sub>:FiO<sub>2</sub>, ratio of partial pressure of arterial oxygen (PaO<sub>2</sub> in mmHg) to fractional inspired oxygen (FiO<sub>2</sub>); COPD, chronic obstructive pulmonary disease; OSAS/OHS, obstructive sleep apnea syndrome/obesity-hypoventilation syndrome; LDH, lactate dehydrogenase.
<sup>†</sup> Missing data: 35 (38.9) Methylprednisolone, 26 (31.3) control group;
<sup>□</sup> Missing data: 26 (28.9) Methylprednisolone, 9 (10.8) control group;
<sup>§</sup> Missing data: 15 (16.7) Methylprednisolone, 17 (20.5) control group;
°P-value of the Fisher's exact test for dichotomous variables, unpaired Student's t-test or Wilcoxon. rank-sum test for numerical variables, as appropriate.

**Table 2:** Distribution of 173 study patients according to study group and clinical outcomes at 28 days.

|  | Methylprednisolone<br>(N=83) | Control<br>(N=90) | p-value* | Crude HR<sup>°</sup><br>(95% CI) | Adj. HR<br>(95% CI)<sup>§</sup> | p-value<sup>§</sup> |
| --- | --- | --- | --- | --- | --- | --- |
| **Major outcomes** |  |  |  |  |  |  |
| Composite primary endpoint, no. (%) | 19 (22.9) | 40 (44.4) | 0.003 | 0.43 (0.25-0.74) | 0.41 (0.24-0.72) | 0.002 |
| Transfer to intensive care unit, no. (%) | 15 (18.1) | 27 (30.0) | 0.067 | .. | .. | .. |
| Invasive mechanical ventilation, no. (%) | 14 (16.9) | 26 (28.9) | 0.095 | .. | .. | .. |
| Death, no. (%) | 6 (7.2) | 21 (23.3) | 0.005 | 0.28 (0.11-0.68) | 0.29 (0.12-0.73) | 0.009 |
| **Other outcomes** |  |  |  |  |  |  |
| Mechanical ventilation-free days, mean (SD) | 19.1 (8.7) | 14.3 (11.7) | 0.003 |  |  |  |
| Invasive mechanical ventilation-free days, mean (SD) | 24.0 (9.0) | 17.5 (12.8) | 0.001 |  |  |  |
| Invasive mechanical ventilation, days, median (IQR)<sup>¶</sup> | 7 (5.5 to 15.5) | 14.5 (12 to 22) | 0.031 |  |  |  |
| Required tracheotomy, no. (%) | 0 (0.0) | 12 (13.3) | <0.001 |  |  |  |
| Intra-patient difference between: |  |  |  |  |  |  |
| CRP at day 3 vs baseline, median (IQR) | -85.0 (-133.0 to -42.5) | -22.0 (-65.0 to 21.3) | <0.001 |  |  |  |
| CRP at day 7 vs baseline, median (IQR) | -99.4 (-162 to -62.3) | -66.1 (-116 to -0.7) | <0.001 |  |  |  |
| PaO<sub>2</sub>:FiO<sub>2</sub> at day 3 vs baseline, median (IQR) | 54.0 (7.0 to 155.0) | 6.9 (-41.5 to 77.0) | <0.001 |  |  |  |
| PaO<sub>2</sub>:FiO<sub>2</sub> at day 7 vs baseline, median (IQR) | 97.5 (42.0 to 162.0) | 68.0 (-5.5 to 139.0) | 0.09 |  |  |  |
| Lymphocytes at day 3 vs baseline, median (IQR) | -45 (-285 to 150) | 0 (-110 to 170) | 0.18 |  |  |  |
| Lymphocytes at day 7 vs baseline, median (IQR) | 110 (-170 to 480) | 130 (-140 to 350) | 0.88 |  |  |  |
| Lymphocytes at day 14 vs baseline, median (IQR) | 590 (-70 to 1390) | 600 (230 to 800) | 0.68 |  |  |  |

Legend: HR, hazard ratio; CI, confidence interval; SD, standard deviation; IQR, inter-quartile range; CRP, C-reactive protein; PaO<sub>2</sub>:FiO<sub>2</sub>, ratio of partial pressure of arterial oxygen (PaO<sub>2</sub> in mmHg) to fractional inspired oxygen (FiO<sub>2</sub>).
\*P-value of Chi-square or Fisher's exact test for dichotomous variables, unpaired T-test or Wilcoxon rank-sum test for numerical variables as appropriate.
<sup>°</sup>HR of event among methylprednisolone vs. control group, estimated using Cox-regression model. The crude odds ratio and 95% CI for the composite outcomes is 0.37 (0.19-0.71).
<sup>§</sup> Cox-regression model was adjusted for sex, age, baseline SOFA score, baseline PaO<sub>2</sub>:FiO<sub>2</sub>, and baseline CRP.
<sup>¶</sup> Only ventilated patients

MP-treated patients had a 28 day lower risk of all-cause death than untreated ones (6 deaths, 7.2% vs. 21 deaths, 23.3%; p=0.005), with a corresponding adjusted HR equal to 0.29 (95% CI: 0.12-0.73), indicating a 71% reduction in the risk of death in MP patients compared to controls. The Kaplan-Meier survival curves (**Figure 2**) showed statistically significant difference between groups (log-rank test p=0.003), with survival probabilities at 28 days of 91.6% (95% CI 82.2 - 96.2) for MP treated and 68.2% (95% CI 53.8 – 78.9) for control patients. The HRs did not substantially change when other variables were included in the adjusted Cox-models (e.g. other allowed treatments, BMI, smoking, NPPV, and high-flow nasal cannula, data not shown). The Kaplan-Meier curves shown in **Figure S1** illustrate timing to removal of mechanical ventilation in both groups.

**Figure 2.**
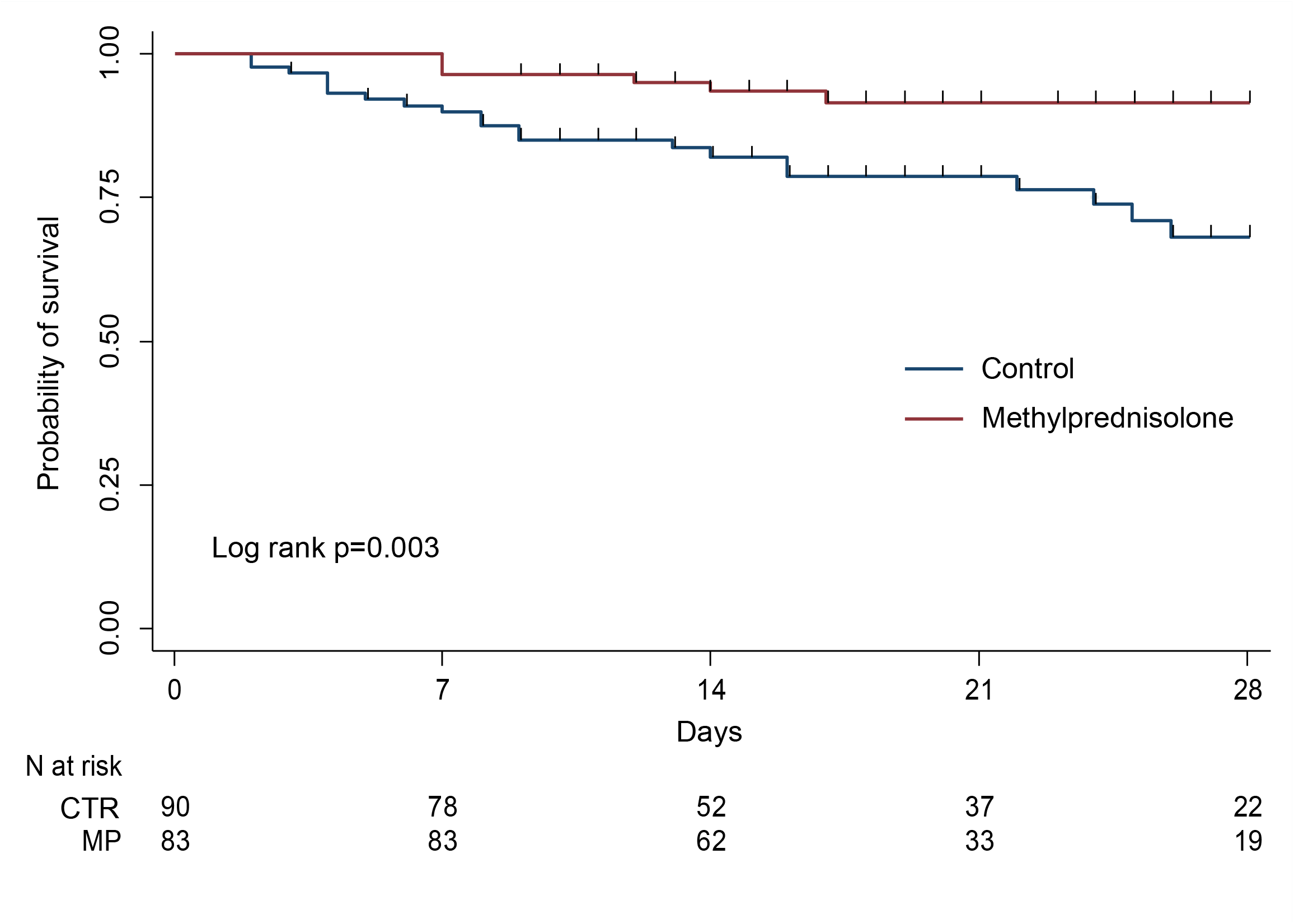
Kaplan-Meier estimates of survival probability. MP, Methylprednisolone; CTR, control.

For the secondary endpoints (**Table 2**), we observed a significant increment in both MV-free days by day 28 outcomes, combined invasive MV and NPPV (19.1 ± 8.7 vs. 14.3 ± 11.7, p=0.003), and invasive-MV-free days alone (24 ± 9 vs. 17.5 ± 12.8, p=0.001). MP exposure was associated with a significant intra-patient median variation in a PaO_2_:FiO_2_ at day 3 compared to baseline [54.0 (7.0 to 155.0) vs. 6.9 (−41.5 to 77.0); p<0.001], but not at days 7, 14 and 28 (**Figure 3**). Median variation in CRP levels was also prominent in the MP group at day 3 [−85.0 (−133.0 to −42.5) vs. −22.0 (−65.0 to 21.3); p<0.001] and at day 7 [−99.4 (−162.0 to −62.3) vs. −66.1 (−116.0 to −0.7); p<0.001] compared to baseline, but not at days 14 and 28 (**Figure 3**). No differences were noted between groups in intra-patient median lymphocytes variation at days 3, 7 and 14 compared to baseline, as detailed in **Table 2** and shown in **Figure 3**. The hospital length of stay did not differ between the groups (p-value=0.38). No tracheostomy was necessary in MP patients vs. 12 controls (OR 0.04, 95% CI 0.002 to 0.64, p-value <0.001). Concerning intra-hospital adverse events of any type (**Table S1**) only the occurrence of hyperglycemia in non-diabetic patients, or severe glycemic decompensation in diabetic patients, and agitation was significantly higher in the MP group compared to control (8 vs. 0, p=0.002 and 9 vs. 2, p=0.03 respectively). No adverse event led to MP discontinuation. Concomitant in-hospital treatments are summarized in **Table S2**.

**Figure 3.**
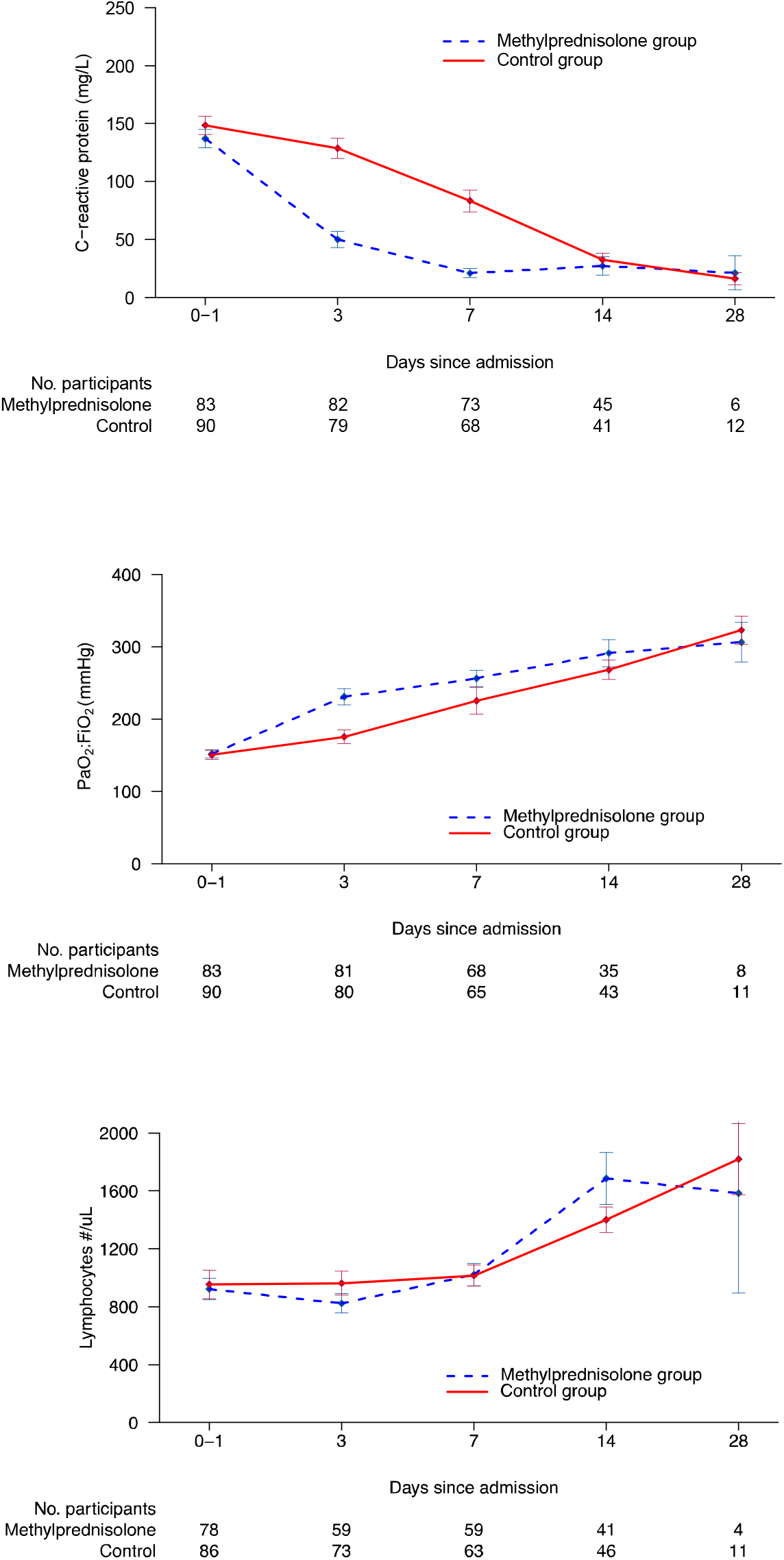
Time-course of C-reactive protein and PaO_2_>:FiO_2_ variation. Upper panel: time-course of C-reactive protein levels (mean ± standard error). The differences between groups were significant at days 3 and 7. Middle panel: time course of mean PaO_2_:FiO_2_. The differences between groups was significant at day 3. Lower panel: time course of mean lymphocyte count showing no significant differences between groups.

There were no relevant differences in viral genome sequencing in the two first recruited patients compared to the average sequences reported in open-source repositories (**Figure S2**). Nor was any observed in viral shedding, determined as time lapse (days) between hospital admission and the first negative RT-PCR for SARS-CoV-2 nasopharyngeal swabs, in a sample of 41 MP-treated patients compared to 28 untreated ones (19.05 ± 6.11 vs. 20.68 ± 7.05, p-value=0.31).

Sensitivity analysis (**Table S3**) show that the primary composite outcome still significantly differs between the MP and control group in scenarios biased against the original hypothesis.

## Discussion

The World Health Organization (WHO) urged the scientific community to carry out urgent evaluation of corticosteroid treatment in patients with severe COVID-19 pneumonia.[13] In our multicenter study, patients exposed to MP encountered the primary composite endpoint of ICU referral, need for invasive MV or in-hospital all-cause death significantly less compared to the control group (adjusted HR 0.41). By day 28, MP treatment was associated with a significant reduction in mortality (adjusted HR 0.29) and an increase in MV-free days. Among patients transferred to the ICU, MP treated patients had a 7.5 days median reduction (p=0.03) in the duration of invasive MV. In line with this data, fewer MP-treated patients required tracheotomy than controls (0 vs. 12, p <0.001). MP-treated patients had a higher reduction in CRP levels than controls. This was statistically significant on days 3 and 7 from baseline and there was a quicker improvement in PaO_2_:FiO_2_ ratio on day 3 for MP-treated patients. There was no overall increase in adverse events between groups, except for an increase in hyperglycemia and mild agitation in the MP-treated patients; no adverse event necessitated MP discontinuation. No difference was observed in viral shedding, determined as the number of days between hospital referral and the first negative nasopharyngeal swab.

Early interventions aimed at down regulating the SARS-CoV-2-associated hyper-immune response in severe COVID-19 patients may well avoid disease progression and enhance pneumonia resolution. The cytokine profile reported for these patients[14] is within the broad range of regulation provided by corticosteroids,[15] particularly MP that is associated with an optimal lung penetration.[16] Our study protocol involved an initial IV bolus to achieve rapid, almost complete glucocorticoid receptor saturation, followed by an infusion to reach a total 160-milligram dose over the first 24 hours. This might explain the rapid reduction observed in inflammatory markers. Treatment duration was guided by monitoring the anti-inflammatory response and oxygenation after at least 7 days. Our MP treatment response is similar to that of RCTs in non-viral ARDS[17] and large-scale observational studies, in severe pneumonia caused by SARS-CoV (n=7008)[18–20] and H1N1 (n=2141) influenza.[21] Additional support for the use of methylprednisolone in COVID-19 originates from transcriptomics data. After matching the expression changes induced by SARS-CoV2 in human lung tissue tissues and A549 lung cell line against the expression changes triggered by 5,694 FDA-approved drugs, methylprednisolone was found to be the drug with the greatest potential to revert the changes induced by COVID-19.[22]

The safety profile reported in our study is consistent with the findings of multiple RCTs investigating prolonged corticosteroid treatment in thousands of patients with severe sepsis, septic shock and ARDS.[17] In these RCTs, hyperglycemia was transient in response to the initial loading bolus and did not impact negatively on outcome.[17] Viral shedding in both groups of our study was in agreement with international literature.[23, 24] Moreover, there is no evidence linking delayed viral clearance to worsened outcome in critically ill COVID-19 patients, and it is unlikely that it would have a greater negative impact than the host’s own cytokine storm.[25]

The observational design of our study implies some obvious limitations, namely a possible restricted control over data collection and potential inclusion biases. However, internal validity was achieved by (1) the comparability of concurrent groups at baseline, (2). accounting for potential confounders into the multivariable Cox regression analyses, and (3) conducting sensitivity analysis to assess for potential bias in outcome ascertainment potentially influenced by medical decision making. Our study’s strengths include a prospective evaluation of a pre-designed intervention protocol based on established pharmacological principles in patients at high risk of progression to ARF and death. Limitations of the study is that we did not control for center effects and site investigators were not blinded to treatment as with any observational study. Despite these limitations, we believe that our findings represent valid and generalizable conclusions, and evidence will be further strengthened with a RCT.

A RCT would provide the most robust evidence on the efficacy of corticosteroid treatment in COVID-19 pneumonia; however, a rapidly developing pandemic and saturated health care systems create serious constraints. This is demonstrated by the lack of RCTs on corticosteroid treatment in prior large respiratory viral epidemics (SARS-CoV-2, MERS-CoV, H1N1). Even so, in the absence of RCTs, well designed prospective observational studies can provide clinicians with valuable data on safety and efficacy. Indeed, we observed benefits when MP treatment was started early and prolonged in COVID-19 pneumonia patients at high risk of ARF progression. MP treatment was demonstrated to be safe, and also allowed for a significant reduction in mortality and immediate improvements in systemic inflammation and oxygenation markers, as well as reducing invasive MV times. We believe our data support the hypothesis that early MP treatment can decrease ICU burden and mortality, therefore reducing the concern surrounding this therapeutic approach in patients admitted with ARF due to severe SARS-CoV-2 pneumonia in the current state of affairs.

## Data Availability

All data referred to in the manuscript are available on reasonable external request.

## Acknowledgments

The authors would like to thank Barbara Wade, contract professor at the University of Torino, for her linguistic advice; Amanda Busby, University of Hertfordshire, for her statistical suggestions; Dr. Valentina Luzzi and Dr. Marco De Martino for their help in data collection.

